# Divergent inflammatory and neurology-related plasma protein profiles in individuals with long COVID following primary and breakthrough SARS-CoV-2 infections

**DOI:** 10.1101/2024.09.06.24312838

**Authors:** Amit Bansal, Sam W.Z. Olechnowicz, Nicholas Kiernan-Walker, Jacob Cumming, Ramin Mazhari, COVID PROFILE consortium, Rebecca J. Cox, Ivo Mueller, Rory Bowden, Emily M. Eriksson

**Affiliations:** Influenza Centre, Department of Clinical Science, University of Bergen, Bergen, Norway; Department of Infectious Diseases, The University of Melbourne, at the Peter Doherty Institute for Infection and Immunity, Melbourne, Australia; The Walter and Eliza Hall Institute of Medical Research, Advanced Technology and Biology Division, Melbourne, Australia; The University of Melbourne, Department of Medical Biology, Melbourne, Australia; The Walter and Eliza Hall Institute of Medical Research, Population Health and Immunity Division, Melbourne, Australia; Disease Elimination Program, Burnet Institute, Melbourne, Australia; The University of Melbourne, Department of Mathematics and Statistics, Melbourne, Australia; Department of Microbiology, Haukeland University Hospital, Bergen, Norway

## Abstract

Long COVID is a complex condition where symptoms persist for more than 3 months after SARS-CoV-2 infection and affects an estimated 5-30% of individuals. While the pathobiology of long COVID is still evolving, persistent inflammation has emerged as an important feature of this condition. However, it is unclear if immune responses from COVID-19 vaccination or SARS-CoV-2 re-infection exacerbate or mirror the initial inflammatory responses. To address this question, we quantified 182 inflammatory and neurology-related proteins in plasma using multiplexed affinity proteomics. Plasma samples were collected 6-9 months after first infection, but before COVID-19 vaccination from individuals who had recovered from COVID-19 (n=21) or from individuals with long COVID (n=12). This was benchmarked against plasma from unvaccinated, SARS-CoV-2 naive individuals (n=24). In addition to this cross-sectional analysis, we performed longitudinal analysis in a subset of individuals (n=34), where paired samples collected 2-4 weeks after a third COVID-19 vaccine dose and after SARS-CoV-2 breakthrough infection were available. Boruta feature selection and lasso regression models identified a distinct plasma profile in long COVID individuals, characterised by elevated IL-20, HAGH, NAAA, CLEC10A, LXN, and MCP-1 and reduced TRAIL, G-CSF, NBL1, and CCL23 protein concentrations. Notably, longitudinal analysis demonstrated that neither COVID-19 booster vaccination nor breakthrough infection replicated inflammatory and neurology-related plasma protein profiles observed after primary infection suggesting an altered immune response outcome in individuals with long COVID upon re-exposure.

## Introduction

After initial SARS-CoV-2 infection, a significant proportion of individuals develop persistent systemic symptoms that may continue for months. This condition is commonly referred to as long COVID or post-acute sequelae of COVID-19 ^1,2^. One of the major challenges in the field is the insufficient understanding of the underlying pathophysiology of this condition, coupled with the absence of reliable biomarkers that could be used in standardised diagnostic tools. Currently, the diagnosis of long COVID is predominantly based on clinical evaluation. The lack of standardised molecular and quantitative diagnostic criteria has impeded accurate estimation of long COVID prevalence. However, available estimates suggest that long COVID affects 10–30% of non-hospitalised individuals and as many as 50–70% of those who were hospitalised during acute infection ^2,3^. Although COVID-19 vaccination may reduce the risk, the incidence of developing long COVID post-vaccination is still estimated to be 10–12% ^2,4,5^.

While the precise mechanisms underlying long COVID is poorly understood, distinct patterns of immune activation have been observed in affected patients and persist several months after SARS-CoV-2 infection. These patterns are evident both at the cellular level and through changes in soluble factors in the bloodstream. The latter includes elevated or altered concentrations of circulating inflammatory mediators^6–9^, complement proteins ^10,11^, and neuro-inflammatory molecules ^9,12^ and inflammatory markers have been associated with a range of persistent long COVID symptoms ^13^. Collectively, these studies have provided essential insights into dysregulated processes that may underlie long COVID. However, many of these long COVID studies have been conducted in populations with high community transmission of SARS-CoV-2 raising concerns about the potential confounding effects of ongoing viral exposure(s) particularly on inflammatory biomarker measurements. Moreover, studies that have monitored inflammatory and neurology plasma protein profiles longitudinally in long COVID individuals remain limited.

In this study, we leveraged access to samples collected from an Australian population with low community transmission during the two first years of the COVID-19 pandemic ^14^. Using a multiplexed affinity proteomics approach, the Olink Target Proximity-Extension Assay (PEA, Olink) and longitudinal plasma samples from initially SARS-CoV-2 naïve individuals, COVID-19 recovered and long COVID individuals, we explored inflammatory and neurology-related plasma protein profiles not only following a primary infection with the SARS-CoV-2 ancestral strain, but also after COVID-19 booster vaccination and breakthrough infections. Our study design provided a rare opportunity to understand how circulating protein profiles after vaccination and re-exposure in the same individual compare to plasma profiles after primary SARS-CoV-2 infections in long COVID individuals.

We identified 14 top inflammatory and neurology-related proteins that were distinctively associated with either long COVID or recovery. These included neuroblastoma suppressor of tumorigenicity 1 (NBL1), interleukin (IL)-20, latexin (LXN), monocyte chemoattractant protein 1 (MCP-1, CCL2), C-type lectin domain containing 10A (CLEC10A), chemokine ligand 23 (CCL23), TNF-related apoptosis-inducing ligand (TRAIL), hydroxyacylglutathione hydrolase (HAGH), granulocyte-colony stimulating factor (G-CSF), N-acylethanolamine-hydrolysing acid amidase (NAAA), artemin (ARTN), cystatin-D (CST5), fibroblast growth factor 19 (FGF-19), and delta and notch-like epidermal growth factor-related receptor (DNER). Notably, long COVID patients exhibited plasma profiles with atypical levels of these proteins, suggestive of ongoing dysregulated processes 6-9 months post-infection. Longitudinal analysis revealed partial resolution of elevated plasma protein levels following COVID-19 booster vaccination, but notably, inflammatory and neurology-related protein patterns observed after primary infection were not replicated following a subsequent breakthrough infection. Our analyses provide pivotal understanding of vaccine-associated and subsequent SARS-CoV-2 infection-induced responses in long COVID individuals and suggests that there is an altered immune response outcome in these individuals upon re-infection without features of dysregulation.

## Methods

### COVID PROFILE study cohort and samples

Plasma was collected as part of COVID PROFILE, a community-based longitudinal cohort study conducted in Victoria, Australia from October 2020 to April 2023^14^. The study was approved by the Walter and Eliza Hall Institute (#20/08) and Melbourne Health (RMH69108) Human Research Ethics Committees. All subjects provided written informed consent to participate in the study, in accordance with the Declaration of Helsinki. Participants were enrolled based on SARS-CoV-2 infection status and initially categorised as COVID-19 recovered or SARS-CoV-2 naïve (confirmed by SARS-CoV-2 PCR/serology and referred to as healthy). Demographic data and detailed clinical information on COVID-19 symptoms, comorbidities, vaccination and breakthrough infections were collected at enrolment and during follow-up visits. Primary vaccination consisted of either two doses of the BNT162b2 mRNA vaccine (COMIRNATY, Pfizer/BioNTech), or ChAdOx1 adenoviral vectored vaccine (Oxford-AstraZeneca), followed by a third mRNA vaccine consisting of either BNT162b2 or mRNA-1273 (Moderna Tx Spikevax). Due to emergence of the long COVID condition, additional follow-up surveys were conducted to identify participants that were experiencing long COVID. These surveys collected comprehensive information on symptom duration and individuals with long COVID were identified in accordance with WHO definition: symptoms persisting or new symptoms 3 months after the initial SARS-CoV-2 infection, with these symptoms lasting for at least 2 months with no other explanation^1^. All other participants experienced COVID-19 symptoms that lasted <90 days post-diagnosis and were classified as COVID-19 recovered. Plasma collected at enrolment for healthy (n = 24) and 6-9 months after primary COVID-19 diagnosis for COVID-19 recovered (n = 21) and long COVID individuals (n = 12) were used for cross-sectional analyses. In a subset of individuals (n=34), additional paired samples collected at 2-4 weeks after a third dose of COVID-19 vaccine and post-breakthrough SARS-CoV-2 infection were used for analysis and only individuals with all three timepoints were included in the longitudinal analysis. Participant demographic data for both analyses are summarised in Table 1.

**Table 1:** Baseline demographic and clinical characteristics of the study cohort

| Variable | Baseline (N = 57) |  |  |  | Longitudinal (n = 34) |  |  |  |
| --- | --- | --- | --- | --- | --- | --- | --- | --- |
|  | Healthy*<br>N = 24<sup>1</sup> | Recovered<br>N = 21<sup>1</sup> | Long<br>COVID<br>N = 12<sup>1</sup> | p-value<sup>2</sup> | Healthy<br>N = 13<sup>1</sup> | Recovered<br>N = 13<sup>1</sup> | Long<br>COVID<br>N = 8<sup>1</sup> | p-value<sup>2</sup> |
| Sex: Female | 14 / 24<br>(58%) | 17 / 21<br>(81%) | 7 / 12<br>(58%) | 0.2 | 9 / 13<br>(69%) | 12 / 13<br>(92%) | 4 / 8<br>(50%) | 0.11 |
| Age: Median<br>(IQR) | 37 (30,<br>46) | 43 (36, 57) | 56 (43,<br>60) | 0.035 | 44 (37,<br>47) | 49 (41, 57) | 57 (53,<br>60) | 0.068 |
| Age range<br>(min-max) | 20, 68 | 21, 66 | 30, 67 |  | 20, 63 | 21, 66 | 30, 66 |  |
| Comorbidity<sup>#</sup> | 3 / 24<br>(13%) | 10 / 21<br>(48%) | 7 / 12<br>(58%) | 0.008 | 3 / 13<br>(23%) | 6 / 13<br>(46%) | 7 / 8<br>(88%) | 0.019 |
| COVID-19<br>disease status |  |  |  | <0.001 |  |  |  | <0.001 |
| Healthy or<br>uninfected* | 24 / 24<br>(100%) | 0 / 21 (0%) | 0 / 12<br>(0%) |  | 13 / 13<br>(100%) | 0 / 13 (0%) | 0 / 8<br>(0%) |  |
| Asymptomatic | - | 1 / 21<br>(4.8%) | 0 / 12<br>(0%) |  | - | 1 / 13<br>(7.7%) | 0 / 8<br>(0%) |  |
| Mild-Moderate | - | 18 / 21<br>(86%) | 12 / 12<br>(100%) |  | - | 11 / 13<br>(85%) | 8 / 8<br>(100%) |  |
| Severe | - | 2 / 21<br>(9.5%) | 0 / 12<br>(0%) |  | - | 1 / 13<br>(7.7%) | 0 / 8<br>(0%) |  |
| Vaccine:<br>Two-dose<br>schedule |  |  |  | 0.2 |  |  |  | 0.3 |
| AstraZeneca | 5 / 24<br>(21%) | 3 / 21<br>(14%) | 5 / 12<br>(42%) |  | 4 / 13<br>(31%) | 2 / 13<br>(15%) | 4 / 8<br>(50%) |  |
| Pfizer | 19 / 24<br>(79%) | 18 / 21<br>(86%) | 7 / 12<br>(58%) |  | 9 / 13<br>(69%) | 11 / 13<br>(85%) | 4 / 8<br>(50%) |  |
| Vaccine:<br>Third dose |  |  |  |  |  |  |  | >0.9 |
| Moderna | - | - | - | - | 3 / 13<br>(23%) | 4 / 13<br>(31%) | 2 / 8<br>(25%) |  |
| Pfizer | - | - | - | - | 10 / 13<br>(77%) | 9 / 13<br>(69%) | 6 / 8<br>(75%) |  |

<sup>1</sup>n / N (%)
<sup>2</sup>Fisher's exact test; Kruskal-Wallis rank sum test
- Not relevant

### Luminex Multiplex assay

A SARS-CoV-2 multi-antigen serology assay was used to detect SARS-CoV-2-specific IgG antibodies against Spike and nucleoprotein (NP) antigens as described previously ^15^. Briefly, fluorescent and magnetic beads were coated with antigens of interest and incubated with individual plasma samples to allow binding of antigen-specific antibodies. Bound antibodies were detected using a secondary anti-human IgG fragment crystallisable region (Fc) antibody conjugated to phycoerythrin (PE) and analysed using MAGPIX^®^ system. The levels of antigen-specific antibodies were reported as mean fluorescent intensity (MFI).

### Affinity proteomics assays

Protein biomarkers were quantified using the Proximity Extension Assay at the WEHI Genomics facility, on the Olink Target platform using Inflammation and Neurology panels (Olink Proteomics, Sweden) for a total of 182 protein targets. Briefly, 1µL of each plasma sample per panel was incubated with a multiplexed set of two oligo-labelled detection antibodies per protein target, which after proximity extension can form a specific DNA barcode ^16^. The barcodes were pre-amplified then detected by qPCR using 96.96 Protein Expression integrated fluidic circuits (IFCs) on a BioMark HD with HX controller (Fluidigm / Standard BioTools, USA). Detection cycle threshold or C(t) values were then processed to NPX values by normalising to the internal Extension Control and the external Inter-plate Control using Olink^®^ Signature software, and batch correction was applied by intensity normalisation and the OlinkAnalyse R package^17^. The values are expressed in arbitrary NPX units that are specific to Olink Proteomics platform and are on a log_2_x scale.

### Statistical analyses

Participant were stratified according to SARS-CoV-2 infection and COVID-19 booster status. Statistical analyses were conducted in R version 4.3.2 (2023-10-31) for macOS using the lme4, ggplot2, ggeffects, ggthemes, patchwork, OlinkAnalyze, performance, Boruta, glmnet, glmnetUtils rpart, rattle, e1071, corrplot and corrr packages.

Descriptive statistics were used to compare participant demographic, clinical characteristics, and exposure factors (Kruskal–Wallis rank-sum test for continuous or Fisher’s exact test for categorical variables). Correlation matrices were plotted by Spearman’s rank correlation test with hierarchical clustering using corrplot package.

To identify potential protein biomarkers, we used a combination of penalised regression (lasso) and feature selection algorithm, using glmnet and Boruta packages in R. The glmnet fitted a generalised linear model via penalised maximum likelihood (lasso, least absolute shrinkage and selection operator) to enhance the prediction accuracy and interpretability of the resulting statistical model^18,19^. The incorporated model fitted a traditional logistic regression model for the log-odds with function glmnet(x, y, standardise=TRUE, alpha = 1, family = “binomial”, intercept = T); where x variable was standardised prior to fitting the model sequence and alpha=1 for the lasso penalty. The model demonstrated superior performance when employing lasso penalty (alpha=1) compared to a ridge penalty (alpha=0) within the elastic-net regularisation path for regression. Analyte selection also involved the Boruta package in R, which uses wrapper algorithms coupled with random forest classifiers ^20^. The overall accuracy rate is computed along with a 95 percent confidence interval (95% CI) for this rate (using exact binomial test; binom.test function) and unweighted Kappa statistics are also calculated. Both accuracy and kappa range from -1 to 1. A value of 1 indicates perfect agreement, while a value of -1 indicates complete disagreement.

F1 score = (1+ beta^2^)*precision*recall/((beta^2^*precision) + recall), where precision = A/(A+B) and recall = A (A+C).

Kappa = (Observed agreement - Chance agreement) / (1 - Chance agreement) or Kappa = 2* (TP * TN – FN * FP) / (TP + FP) * (FP + TN) + (TP + FN) * (FN + TN)

Sensitivity = A/(A+C)

Specificity = D/(B+D)

Accuracy = (TP + TN) / (TP + TN + FP + FN).

Boxplots were plotted displaying the distribution of data; with minimum value, the first quartile (Q1), the median, the third quartile (Q3), and the maximum value. The olink_anova_posthoc performs a post-hoc ANOVA test using the function emmeans from the R library emmeans with Tukey p-value adjustment per assay (by OlinkID) at confidence level 0.95, p-value annotations: “***”=0.001, “**”=0.01, “*”=0.05.

We employed recursive partitioning and regression trees (rpart) to identify protein analytes predictive of COVID-19 status (long COVID and COVID-19 recovered, long COVID and healthy, and COVID-19 recovered and healthy) before vaccination (n = 57). Models were constructed using rpart and fancyRpartPlot functions in R to understand the hierarchical relationships and thresholds of protein analytes, extracted by lasso and Boruta random forest, that differentiated COVID-19 status (long COVID and COVID-19 recovered, long COVID and healthy, and COVID-19 recovered and healthy). Decision boundaries, generated using support vector machines (SVM) algorithms from the e1071 package in R ^21^, were plotted to distinguish between the groups (healthy, recovered, and long COVID) based on the selected protein concentrations.

Longitudinal data (n = 34) was analysed by olink_lmer function, which fitted a linear mixed-effects model for each protein analyte or antibodies as outcome measure. It uses the lmer function from the R library lmerTest and the anova function from the R library stats. Adjusted p-values are calculated using the p.adjust function from the R library stats with the Benjamini & Hochberg method. The threshold for significance is p < 0.05. The olink_lmer_posthoc function is used to extract and plot significant proteins. It performs a post-hoc analysis based on a linear mixed-effects model using the lmer function. Immune dynamics were plotted using linear mixed-effects models fitted by the lmer function of the lme4 package. The resulting linear equation of the model was y = α + β*Group*Condition +(1 | Participant), where group is disease status and condition are exposure factors. Randomisation was not utilised, as the study aimed to compare individuals with long COVID individuals with those recovered or uninfected or healthy controls. Data collection and analysis were not conducted in a blinded manner with respect to the experimental conditions. Both unsupervised and supervised machine learning algorithms for protein analyte selection were applied, selecting models that demonstrated relatively good fit.

## Results

### Cohort characteristics

A total of 57 participants (aged 20 to 68 years; 38 females, 67%) enrolled in a longitudinal cohort conducted from October 2020 to April 2023 were included in this study. The participants were categorised into three groups: healthy SARS-CoV-2 PCR/serology confirmed uninfected individuals (n = 24; age 20-68 years, median age 37; 14 (58%) females), COVID-19 recovered (n = 21; age 21-66 years, median age 43; 17 (81%) females) and long COVID (n = 12; age 30-67 years, median age 56; 7 (58%) females) with no significant sex difference between groups but median age higher in the long COVID group (p=0.035) (Fig. 1 and Table 1). Most participants experienced mild to moderate primary COVID-19 disease with only 9.5% of individuals of the COVID-19 recovered group requiring hospitalisation due to severe disease (Table 1). A range of co-morbidities (including well-controlled asthma, hypothyroidism, cancer or other chronic illnesses) was observed among the healthy (13%), COVID-19 recovered (48%) and long COVID (48%) participants. Half (50%, 6/12) of individuals diagnosed with long COVID reported an alleviation of symptoms during follow-up (4 became asymptomatic before COVID-19 boosters and 2 before re-infection). Conversely, the remaining cohort (50%) continued to experience long COVID symptomatology both after receiving a booster vaccination and breakthrough SARS-CoV-2 infection. The most common symptom reported in this study cohort was fatigue, brain fog and loss of smell and taste (Fig. 1). Of the 57 participants, longitudinal analysis was performed on 34 participants with paired blood samples at three timepoints: pre-vaccination; 2-4 weeks after receiving a booster COVID-19 vaccine dose; and either after a new SARS-CoV-2 infection (healthy group) or after re-infection (COVID-19 recovered or long COVID groups).

**Figure 1:**
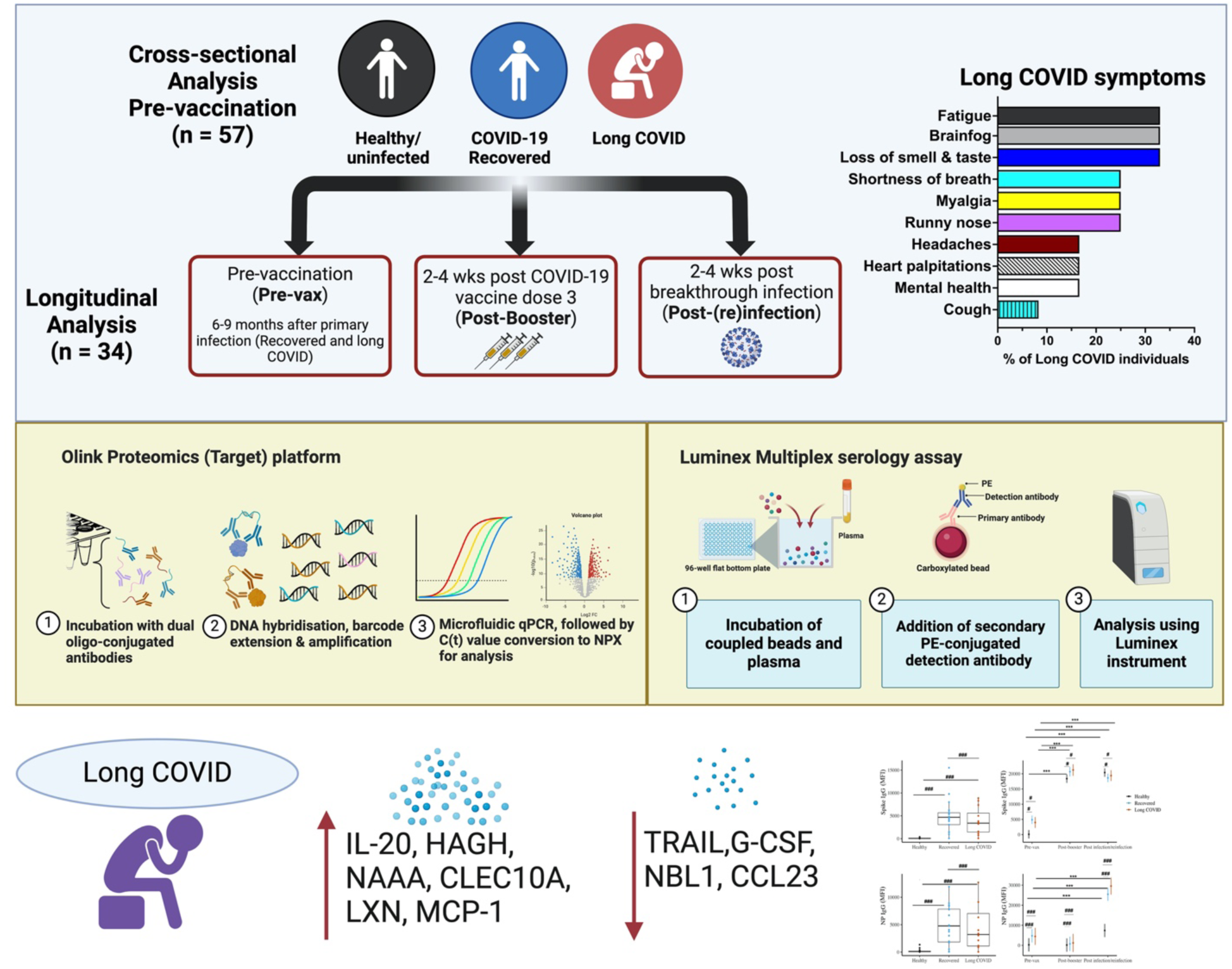
Study overview. Overview of the study including: (Top panel) Cohort participant description, sample collection time points and associated long COVID symptoms. (Middle panel) visual representation of methodology and (Bottom panel) summary of predictive and differentially expressed proteins in long COVID after primary SARS-CoV-2 infection and antibody levels pre-and post-COVID-19 booster vaccination and (re)infection. Created with BioRender.com (2024)

### Distinct patterns of inflammatory and neurology-related proteins in plasma from healthy, COVID-19 recovered individuals and long COVID sufferers

Persistent inflammation has been proposed as a key mechanism underlying long COVID, particularly because it may explain the varied manifestations associated with this condition. To determine whether long COVID individuals in our low transmission cohort align with previous observations of lasting inflammation and whether inflammatory and neurology-related proteins can be predictive of long COVID, we profiled the levels of 182 plasma proteins using multiplexed affinity proteomics. These plasma samples were collected at enrolment for healthy (n = 24) and 6-9 months after primary COVID-19 diagnosis for COVID-19 recovered (n = 21) and long COVID individuals (n = 12). All plasma was collected prior to COVID-19 vaccination for this cross-sectional analysis. Initial analysis mapped the overall plasma protein profiles of each individual (supplementary Fig. S1) and correlations between analytes. as Correlation of proteins associated with long COVID disease status were explored using Spearman’s rank correlation analyses (supplementary Fig. S2 and S3). In this first analysis, only TRAIL was significantly associated with long COVID (r = -0.32, p = 0.014) (supplementary Fig. S3). Correlations of analytes between COVID-19 recovered and long COVID groups (supplementary Fig. S4), healthy and long COVID groups (supplementary Fig. S5) and healthy and COVID-19 recovered groups (supplementary Fig. S6) were visualised using network analyses plots of the 182 analytes, where proteins are grouped into clusters suggesting functional or biological relationships, but not causal interactions. Furthermore, we used gene set enrichment analysis (GSEA) to associate disease status with specific groups of proteins (supplementary Fig. S7). While these GSEA results did not reach statistical significance, the observed pathways highlight the potentially ongoing and altered processes in long COVID at 6-9 months following primary SARS-CoV-2 infection. This also extended to some degree to the COVID-19 recovered group where individuals in the COVID-19 recovered group exhibited upregulated neuroinflammatory pathway responses compared to the healthy group. However, individuals with long COVID displayed a more complex neuroinflammatory response profile compared to the COVID-19 recovered group.

To further understand which analytes differed significantly between healthy, COVID-19 recovered and long COVID groups, lasso regression analysis was also performed. First, we investigated plasma profile differences between long COVID and COVID-19 recovered groups. Lasso regression identified 4 top protein analytes associated with long COVID (NBL1, IL-20, LXN, MCP-1), with an accuracy of 0.55 (95%CI: 0.24-0.83) (Fig 2B and C, Table 2). In addition, Boruta random forest and recursive partitioning and regression trees were conducted to identify proteins predictive of COVID-19 status. Consistent with lasso regression analysis, NBL1 and IL-20 were identified as top features with both Boruta random forest (Fig. 2A and C) with similar accuracy of 0.55 (0.23 - 0.83; Table 2) and recursive partitioning and regression trees (Accuracy = 0.88, 95% CI: 0.72-0.97; and F1 score = 0.91) (Fig 3A and Table 2). IL-20 concentrations were particularly lower in individuals who recovered compared to long COVID individuals (estimate or β = -0.29, 95% CI: -0.49 - -0.09, adj p = 0.007) (Fig. 2C). Conversely, long COVID individuals were more likely to have lower levels of NBL1 (< 4.7) than those who recovered (Figs. 2C, 3A and B).

**Figure 2:**
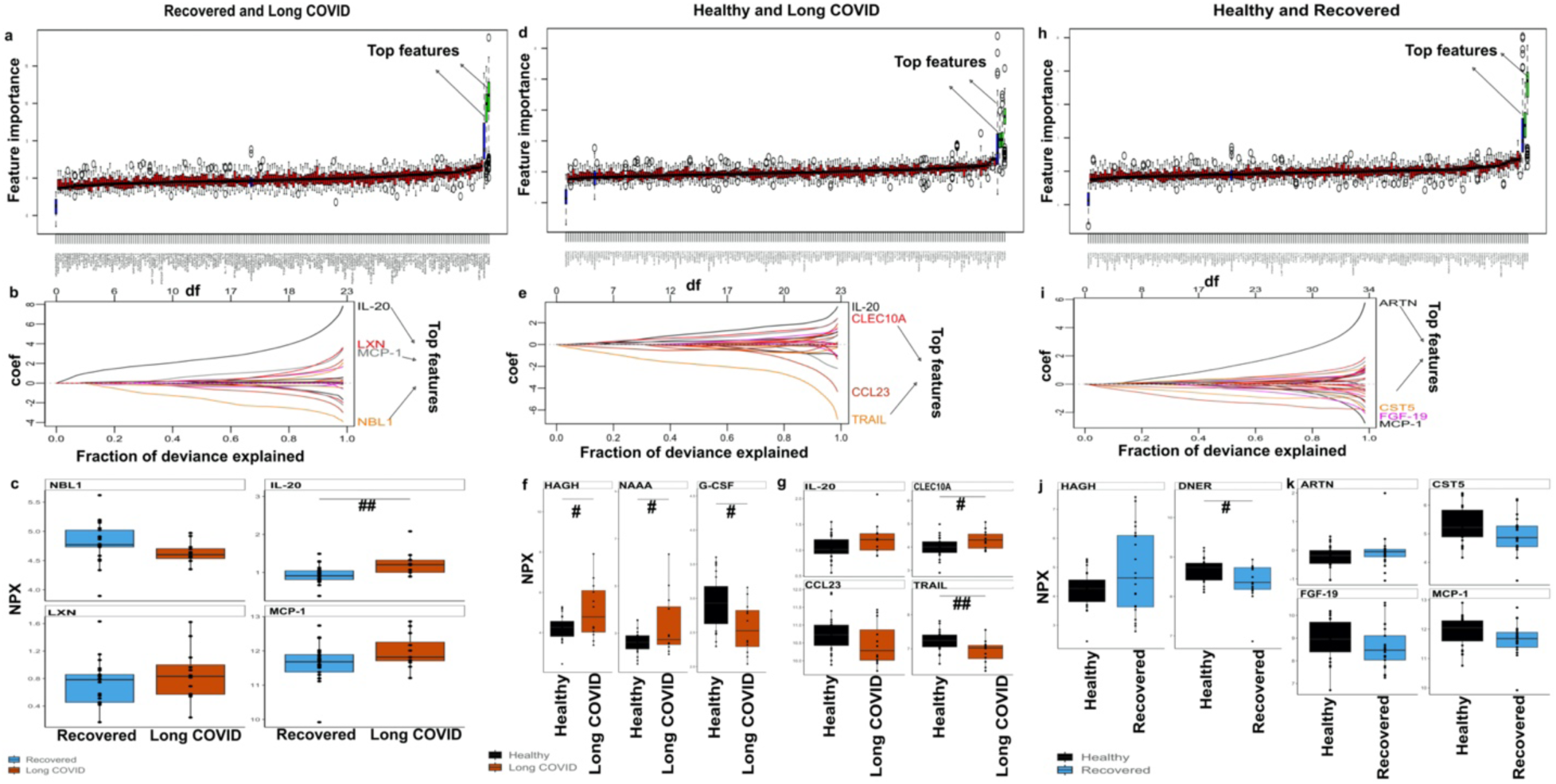
Atypical levels of neurology-related and inflammatory biomarkers in patients with long COVID detected 6-9 months after primary SARS-CoV-2 infection. Data representing plasma levels of 182 proteins from 57 individuals was analysed using Boruta feature selection ^20^ and lasso regression models (glmnet function^18,19^) to identify group differences. Boruta feature selection subplots representing differences between (a) recovered vs. long COVID, (d) healthy vs. long COVID and (h) healthy vs. recovered. Blue box-plots correspond to the shadow attributes (with minimum, mean and maximum attribute values), green box-plots represent important variables, yellow box-plots are tentative, and red box-plots are not significant. Subplots represent the coefficient paths of lasso regression models to identify group differences between (b) COVID-19 recovered vs. long COVID, (e) healthy vs. long COVID and (i) healthy vs. COVID-19 recovered. The x-axis shows the fraction of deviance explained against a coefficient profile plot. Subplots representing boxplots for identified differential proteins between (c) COVID-19 recovered vs. long COVID, (f and g) healthy vs. long COVID and (j and k) healthy vs. COVID-19 recovered individuals. Each circle represents an individual. The lower and upper hinges correspond to the first and third quartiles (25th and 75th percentiles), with the median at the midpoint. Boxplots were created using olink_anova_posthoc and olink_boxplot function ^17^ with Tukey p-value adjustment for multiple testing within an assay at confidence level 0.95; two-tailed p <0.05 #; 0.01-0.001 ##; <0.001 ### were considered significant for group differences.

**Table 2:** Best features identified by selected models at pre-vaccination time point, and models accuracy.

| Condition | No. of features | Best features | Model | F1 score | Kappa | Sensitivity | Specificity | Accuracy(95%CI) |
| --- | --- | --- | --- | --- | --- | --- | --- | --- |
| Recovered vs Long COVID | 4 | NBL1, IL-20, LXN, MCP-1 | LASSO (Least Absolute Shrinkage and Selection Operator) | 0.62 | 0.07 | 0.67 | 0.40 | 0.55 (0.24- 0.83) |
|  | 2 | NBL1, IL-20 | Boruta Random Forest | 0.62 | 0.07 | 0.67 | 0.40 | 0.55 (0.23 - 0.83) |
| Healthy vs Long COVID | 4 | IL-20, CLEC10A, CCL23, TRAIL | LASSO | 0.71 | 0.31 | 0.71 | 0.60 | 0.67 (0.35 - 0.91) |
|  | 3 | HAGH, G-CSF, NAAA | Boruta Random Forest | 0.82 | 0.44 | 0.99 | 0.40 | 0.75 (0.43 - 0.95) |
| Healthy vs Recovered | 4 | ARTN, CST5, FGF-19, MCP-1 | LASSO | 0.67 | 0.18 | 0.99 | 0.20 | 0.56 (0.31- 0.79) |
|  | 2 | HAGH, DNER | Boruta Random Forest | 0.74 | 0.46 | 0.88 | 0.60 | 0.72 (0.47 - 0.90) |

**Figure 3:**
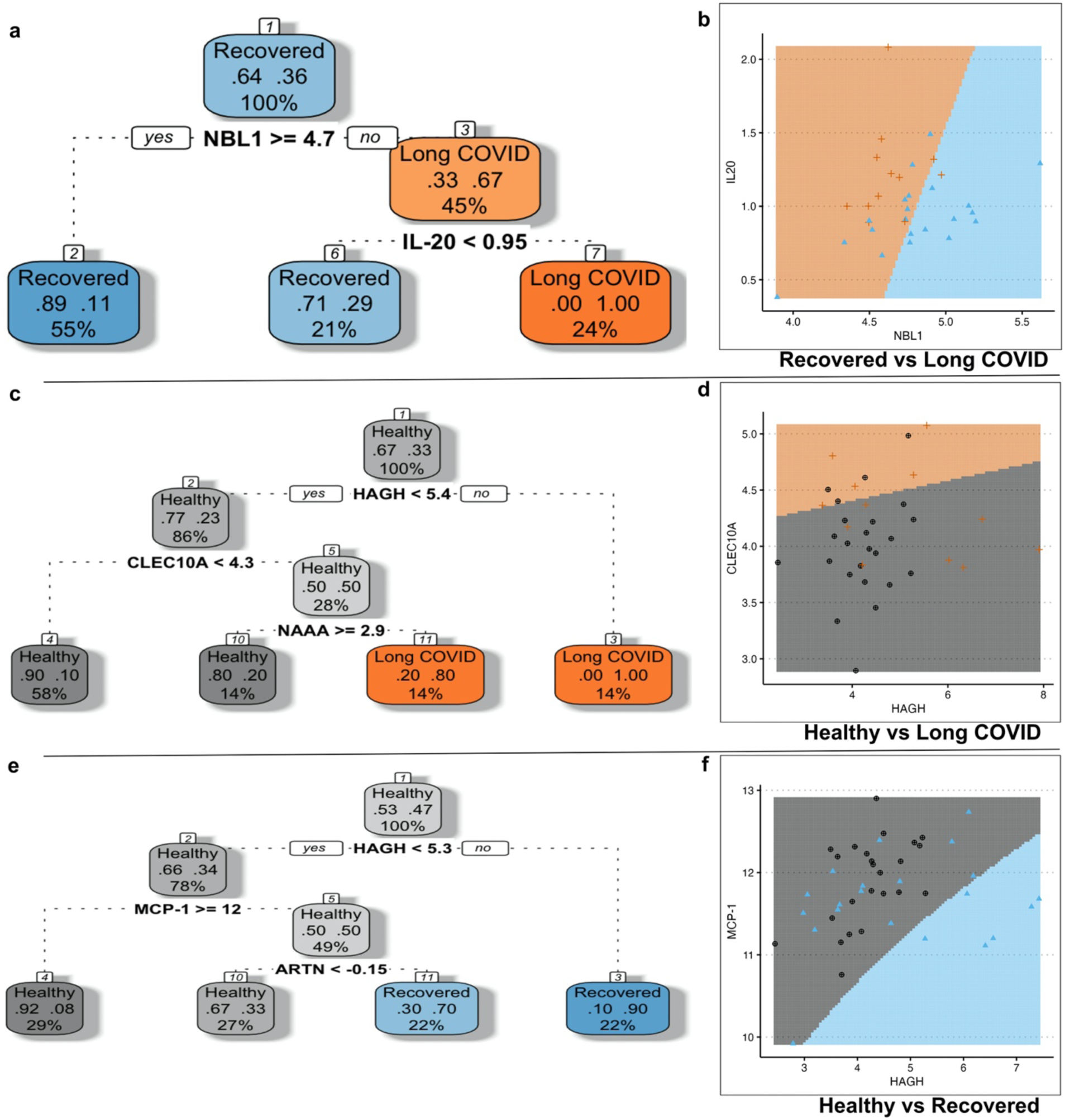
Decision tree and hyperplane boundaries displaying hierarchical relationships and thresholds of protein features detected at 6-9 months after primary SARS-CoV-2 infection to predict long COVID or COVID-19 recovery. Models were constructed using rpart and fancyRpartPlot functions in R to understand the hierarchical relationships and thresholds among protein analytes, extracted by lasso regression models and Boruta random forest models that differentiated between (a) long COVID and COVID-19 recovered, (b) long COVID and healthy, and (c) COVID-19 recovered and healthy ^66,67^. Two-dimensional scatter plots of NPX values for the 2 top protein analytes separating three groups (b) NBL1 and IL-20 for COVID-19 recovered vs. long COVID with 57% accuracy (d) CLEC10A and HAGH for healthy vs. long COVID with 65% accuracy; and (f) HAGH and MCP-1 for healthy vs. COVID-19 recovered with 56% accuracy). A decision boundary, created using support vector machines (SVM) algorithms from the e1071 package in R ^21^, separates the groups based on the protein concentrations.

Using the same approach, analysis of plasma protein profiles between the long COVID and healthy groups identified 4 differential proteins (IL-20, CLEC10A, CCL23, TRAIL) using lasso regression analysis (Fig. 2E) with accuracy of 0.67 (0.35 - 0.91; Table 2), and collectively 5 top predictive proteins (HAGH, G-CSF, NAAA) with Boruta random forest (Fig. 2D, F and G) with accuracy of 0.75 (0.43 - 0.95; Table 2) and (HAGH, CLEC10A and NAAA) using decision tree (Accuracy = 0.89, 95% CI: 0.74-0.97; and F1 score = 0.92; Fig. 3C). The healthy group had lower levels of HAGH (estimate= -0.88, 95% CI: -1.58 - -0.18, adj p = 0.02) and CLEC10A concentrations (β = -0.31, 95% CI: -0.62 - -0.005, adj p = 0.046), but higher TRAIL concentration (β= 0.31, 95% CI = 0.08 - 0.55, adj p = 0.01) compared to individuals with long COVID (Fig 2F and G). Specifically, the decision tree analysis revealed that individuals with HAGH NPX levels > 5.4 and/or CLEC10A >4.3 and NAAA < 2.9 were more likely to be suffering from long COVID compared to the healthy group (Fig. 3C and D).

Finally, we assessed plasma protein profile differences between COVID-19 recovered and healthy groups. In this comparison, a distinct set of differential proteins was identified (ARTN, CST5, FGF-19, MCP-1) using lasso regression (Fig. 2I, J and K) with accuracy of 0.56 (0.31-0.79) (Table 2). Additionally, predictive proteins consisted of HAGH, DNER as determined using Boruta random forest (Fig. 2H) with accuracy of 0.72 (0.47 - 0.90) and HAGH, MCP-1 and ARTN identified using a decision tree (Accuracy = 0.8, 95% CI: 0.65-0.90; and F1 score = 0.82; Fig 3 E and F). The healthy group had higher concentrations of DNER than COVID-19 recovered (β= 0.28, 95% CI: 0.05 - 0.51, adj p = 0.018) and individuals with HAGH NPX levels of > 5.3 and/or MCP-1 <12 and ARTN > -0.15 were more likely to be COVID-19 recovered than healthy (Fig. 3E and F).

Collectively these findings demonstrate that distinct plasma protein profiles can be detected in groups of individuals with different levels of SARS-CoV-2 exposure (healthy vs. previously SARS-CoV-2-infected) and varying disease outcomes (COVID-19 recovered vs. long COVID), even long after SARS-CoV-2 acute infection and diagnosis.

### Lower SARS-CoV-2-specific IgG antibody levels following breakthrough infection in long COVID and COVID-19 recovered individuals compared to healthy individuals

Humoral responses to COVID-19 vaccines and SARS-CoV-2 infections after vaccination (breakthrough infections) result in high levels of antigen-specific antibodies ^22–24^. However, this response is poorly documented in individuals with long COVID. To gain insight into the overall responsiveness to COVID-19 booster vaccination and SARS-CoV-2 breakthrough infection in our cohort, we measured SARS-CoV-2-specific anti-Spike and anti-NP antibody responses in all individuals (n=57) prior to COVID-19 vaccination, but 6-9 months after infection in COVID-19 recovered and long COVID individuals (Fig. 4; left panels). Longitudinal analysis of antibody levels was also performed (Fig. 4; right panels) using paired samples from individuals (n=34) at three timepoints collected at pre-vaccination (pre-vax), 2-4 weeks after the third COVID-19 vaccine dose (post-booster), and 2-4 weeks after either new SARS-CoV-2 infection (healthy group) or being re-infection (COVID-19 recovered or long COVID groups; post(re)infection). As expected prior to vaccination (pre-vax), individuals previously infected with SARS-CoV-2 had significantly higher Spike and NP-specific antibodies than healthy individuals (Fig. 4, p <0.001). However, individuals with long COVID had significantly lower Spike-specific and NP-specific IgG antibodies compared to the COVID-19 recovered group (Fig. 4 left panels, p <0.001) before vaccination. Following booster vaccination all three groups showed high levels of Spike-specific antibodies (Fig. 4; top right panel, p <0.001). At this time point, as expected, NP-specific antibodies remained higher in the COVID-19 recovered and long COVID groups compared to the healthy group (Fig. 4; bottom right panel). After breakthrough infection, all groups had increased NP-specific antibody levels (Fig. 4; bottom right panel). However, vaccinees with long COVID showed substantially higher NP-specific antibodies than newly infected vaccinees (healthy). In contrast, Spike-specific antibodies in the latter group were higher compared to antibody levels measured following re-infection in the COVID-19 recovered and long COVID groups (Fig. 4 top right panel).

**Figure 4.**
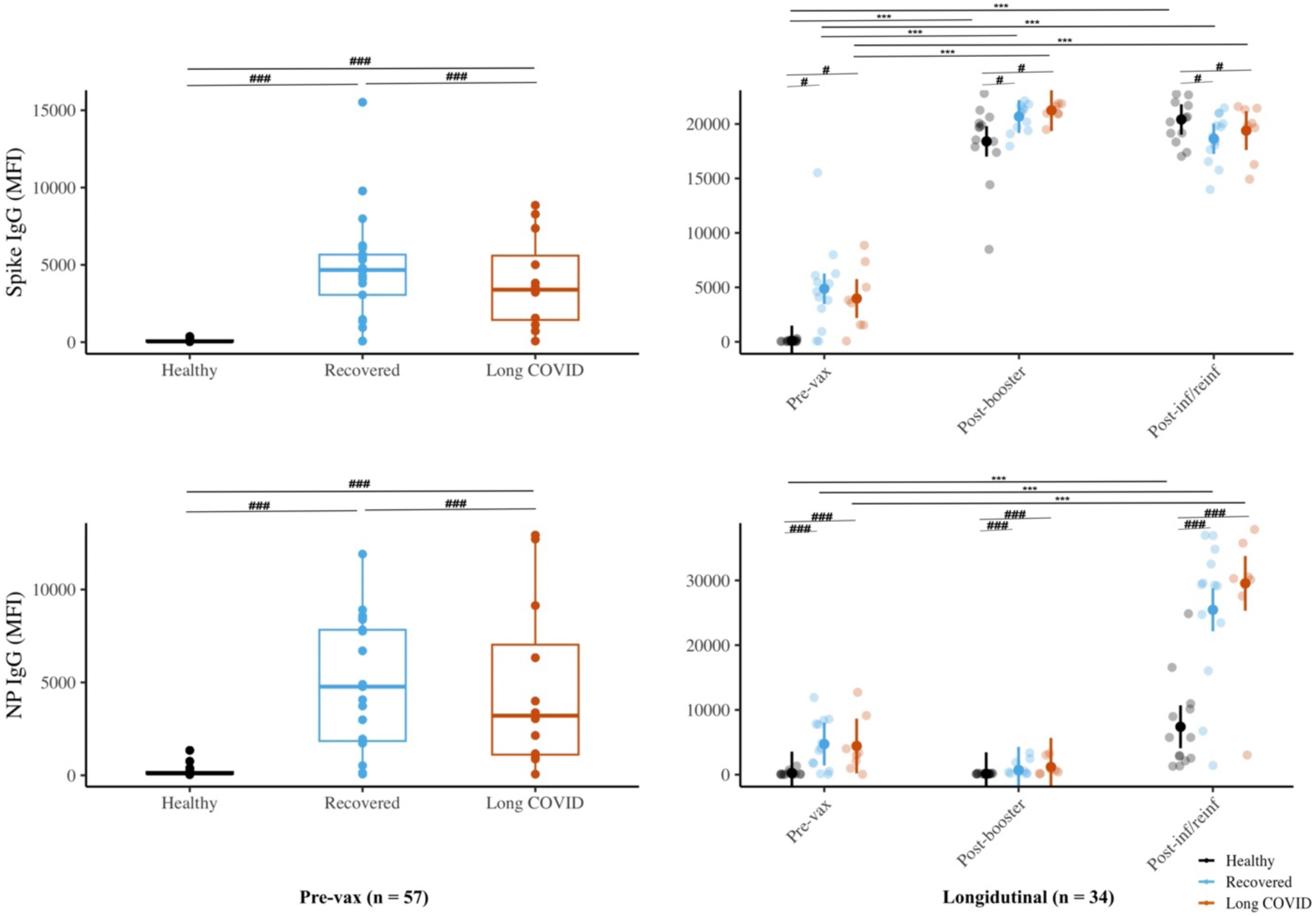
High levels of SARS-CoV-2-specific IgG antibodies post-COVID-19 booster vaccination and post (re)infection. Left panels: Levels of Spike and anti-nucleoprotein (NP) IgG antibodies in healthy (n=24; black), COVID-19 recovered (n=21; blue) and long COVID individuals (n=12; red) 6-9 months after SARS-CoV-2 infection. Each circle represents an individual. The lower and upper hinges correspond to the first and third quartiles (25th and 75th percentiles), with the median at the midpoint. Right panels: Levels of Spike and anti-nucleoprotein (NP) IgG antibodies in healthy (n=13; black), COVID-19 recovered (n=13; blue) and long COVID individuals (n=8; red) 6-9 months after SARS-CoV-2 infection (pre-vax), 2-4 weeks after COVID-19 booster vaccination (post-booster) and 2-4 weeks after SARS-CoV-2 (re)infection (post-(re)infection) The y-axes show antibody levels in median fluorescence intensities (MFI). (Left panels) Wilcoxon Rank Sum and Signed Rank Tests were performed with Tukey p-value adjustment for multiple testing within an assay at confidence level 0.95; two-tailed p <0.05 #; 0.01-0.001 ##; <0.001 ### were considered significant for group differences. (Right panels) Mixed-effects models with the subject as a random effect was performed to analyse longitudinal data from 34 individuals categorised as HC, COVID-19 recovered and long COVID groups with lmer function and Benjamini-Hochberg procedure in R. Estimated marginal means with 95% confidence intervals are depicted as vertical lines with error bars. Individual data points are represented as circles. Two-tailed p <0.05 #; 0.01-0.001 ##; <0.001 ### were considered significant for group differences (recovered vs. long COVID, HC vs. long COVID and HC vs. recovered) and p <0.05 *; 0.01-0.001 **; <0.001 *** for longitudinal changes.

### Differential plasma profiles after COVID-19 booster vaccination, primary and breakthrough infections in long COVID individuals

Vaccines are designed to induce short term inflammatory responses in order to achieve desired immune responses that ultimately provide protection from invading pathogens. However, the impact of vaccine-induced inflammatory responses in individuals that are indicated to experience dysregulated inflammatory activity, such as those with long COVID, is not well-established. Similarly, understanding of inflammatory reactions following breakthrough SARS-CoV-2 infection in long COVID individuals is lacking. To investigate these specific aspects, we analysed longitudinal and paired samples from individuals (n=34) across three timepoints as previously (pre-vaccination, post-booster and post-infection/re-infection). In this mixed-effects models analyses, we extracted 7 significant proteins (LAT, CASP-8, SIRT2, 4E-BP1, CCL4, STAMBP, and HAGH) from the 182 measured inflammatory and neurology-related protein levels using the Olink proteomics assays. These proteins were either different between groups or across time points within a group (Fig. 5). We found that SIRT2 concentrations were higher in the healthy group compared to COVID-19 recovered pre-vaccination (p=0.03; Fig. 5A), and these group differences were maintained following the booster COVID-19 dose (p = 0.04). Similar changes were also observed with 4E-BP1 when comparing healthy and COVID-19 recovered groups (p = 0.02) at pre-vaccination and following booster (p = 0.21; Fig. 5B). Notably, SIRT2 and 4E-BP1 NPX values were similar between the long COVID and healthy groups, both at pre-vaccination and post-booster. However, after breakthrough infection, the levels of these two proteins in both long COVID and healthy groups were significantly lower than at pre-vaccination and aligned with the COVID-19 recovered group. Similar patterns were observed with LAT and STAMBP (Fig 5B and C) although substantial differences were only noted between time points within groups. The protein HAGH, previously identified as a differential and predictive marker at pre-vaccination using Boruta random forest and lasso regression analysis, was not elevated following breakthrough infections in this longitudinal analysis (Fig. 5C). In healthy individuals, CCL4 levels decreased after breakthrough infection, but remained similar for the three sample events in the COVID-19 recovered and long COVID groups (Fig. 5C). Finally, caspase-8 (CASP-8) was significantly higher in the long COVID group at pre-vaccination but decreased to levels similar to those in the COVID-19 recovered and healthy groups. CASP-8 was not induced by either COVID-19 vaccination or a breakthrough infection in any of the three groups (Fig. 5A). Overall, these findings suggest that primary infections and SARS-CoV-2 breakthrough infections result in dissimilar plasma protein profiles, particularly in individuals with long COVID. While COVID-19 booster vaccination is expected to induce a burst of new onset inflammatory responses, this does not appear to exacerbate or persist divergently in long COVID individuals.

**Figure 5.**
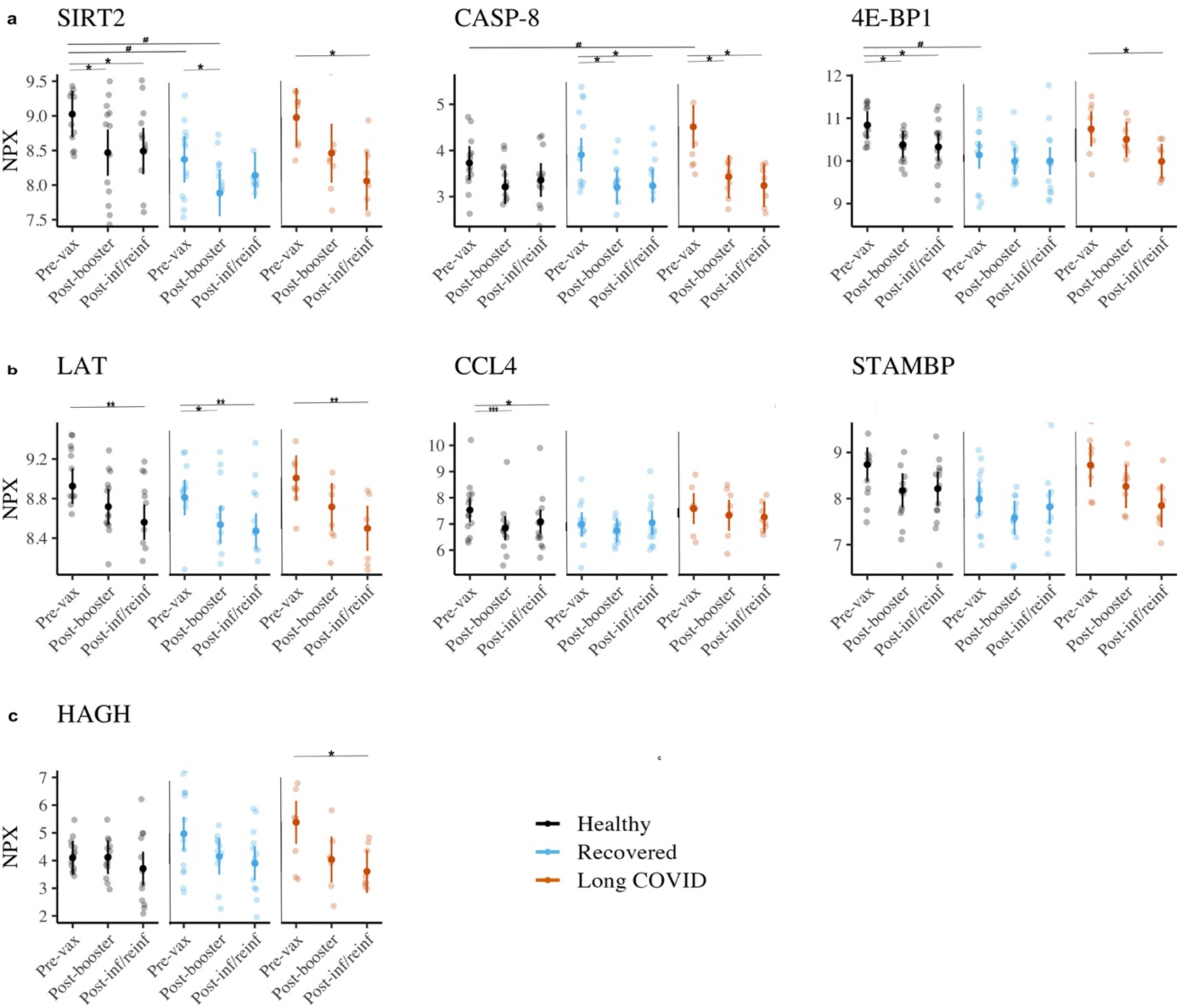
Longitudinal analysis of neuro-inflammatory profiles using mixed-effects models at pre-vaccination, post-COVID-19 booster vaccination and post (re)infections. Mixed-effects models with the subject as a random effect were performed to analyse longitudinal data from 34 individuals categorised as healthy, COVID-19 recovered and long COVID. Paired samples collected at pre-vaccination, 2-4 weeks post COVID-19 booster vaccination and 2-4 weeks post-(re)infection was included for each individual. Linear modelling was performed to identify important proteins and the Benjamini-Hochberg procedure used to control for false discoveries among the 182 assays used ^17^. Within-assay multiple testing correction was applied using the Benjamini-Hochberg procedure with Kenward-Roger degrees-of-freedom method for the 15 selected contrasts. Estimated marginal means with 95% confidence intervals are depicted as vertical lines with error bars. Individual data points are represented as circles. Two-tailed test, p <0.05 #; 0.01-0.001 ##; <0.001 ### were considered significant for group differences (COVID-19 recovered vs. long COVID, healthy vs. long COVID and healthy vs. COVID-19 recovered) and p <0.05 *; 0.01-0.001 **; <0.001 *** for longitudinal changes^68^

## Discussion

Long COVID is a complex condition that can affect multiple organ systems although its pathophysiology remains elusive. Emerging concepts suggest that prolonged inflammation, viral antigen persistence, human herpesvirus reactivation, dysbiosis, microvascular dysfunction and autoreactive immune responses are probable contributing factors^25,26^. In this study we focused on establishing the persistent inflammatory and neurology-related plasma protein profiles after initial disease-related effects subside. We also assessed, primary infections, alongside longitudinal samples to investigate if dysregulated inflammatory reactions are mirrored or exacerbated post-COVID-19 booster vaccination and re-infection. Our results suggest that modified levels of neurology-related and inflammation proteins in plasma can persist up to 6-9 months after primary infection in long COVID individuals. Notably, these primary infection responses are not repeated early after breakthrough infection in long COVID individuals.

We used affinity proteomics to establish inflammatory and neurology-related plasma protein profiles in our cohort sample set. This highly sensitive multiplexed Proximity-Extension Assay (PEA, Olink Proteomics) has increasingly been utilised to both investigate plasma and cerebrospinal fluid (CSF) profiles following COVID-19 in the search for potential biomarkers for long COVID ^12,27–30^. These studies have collectively identified proteins that can be used to classify long COVID, but also show that different markers can be related to specific long COVID subtypes based on symptoms. Similarly, using the same methodology, we identified a panel of plasma proteins from our cross-sectional analysis (NBL1, IL-20, LXN, MCP-1, CLEC10A, CCL23, TRAIL, HAGH, G-CSF, and NAAA) that were either biomarkers of long COVID or observed at differential levels from either healthy or COVID-19 recovered individuals. Of note, decreased levels of TRAIL, a cytokine with anti-inflammatory properties, in CSF was previously associated with long COVID and align with our finding in plasma ^12^. However, while some of our proteins have been reported to be differentially expressed across these specific studies, the remaining proteins in our panel, were not among the proteins previously identified in plasma as predictive of long COVID. It is likely that the divergent panel of proteins reported between the studies is due to the different COVID-19 sub-groups included for comparison. These previous studies focused on differences between acutely ill and/or hospitalised individuals with long COVID sufferers, whereas our study focused on individuals after recovery from mild to moderate disease and long COVID individuals. Importantly our samples were collected within a similar timeframe after diagnosis for both groups. Nevertheless, each study underscores the complexity of long COVID and highlights the importance of considering various comparisons to comprehensively unravel serological features of this condition.

We found that IL-20 was a prominent feature of long COVID in our cohort. IL-20 is a pleiotropic cytokine with potent inflammatory properties and has been positively associated to the severity of SARS-CoV-2 infection ^31^. The observation of increased IL-20 in long COVID individuals is intriguing as IL-20 exerts its effects through the activation of signal transducer and activator of transcription (STAT) proteins and specifically via the type I IL-20 receptor complex, which is composed of IL20RA and IL20RB subunits. Cytokines that use IL-20 receptor complex are believed to be linked to the development of chronic inflammation and autoimmune diseases ^32^ with increased IL-20 concentrations linked to autoantibody production in rheumatoid arthritis patients ^33^. This raises the intriguing possibility that dysregulation of the IL-20 pathway, characterised by enhanced IL-20 levels, might contribute to the development of autoimmune features known to occur in a subset of long COVID individuals ^34,35^. IL-20 also induces synovial fibroblasts and kidney mesangial cells to produce pro-inflammatory molecules, including MCP-1 ^36,37^ a protein that was also found in our study to be a feature of long COVID.

Given that neurological symptoms, including fatigue and cognitive dysfunction such as memory difficulties, attention deficits and sleep disturbances, are common in long COVID and were frequently reported by our patients, we also quantified protein biomarkers with known relation to neurological disease. Interestingly HAGH, LXN, Linker For Activation Of T Cells (LAT) and NAAA have all been related to cognitive performance in older adults with metabolic syndrome ^38^. HAGH is also a potential biomarker of Alzheimer disease ^39^. Increased levels of HAGH have been linked to low cognitive function whereas an inverse relationship of lower levels of LXN, LAT and NAAA were related to higher cognitive function ^38^. Apart from LAT, these were all elevated in the long COVID group in our cross-sectional analysis. However, HAGH does not appear to be an indicator of long COVID as increased HAGH was reported in mild neuro-COVID cases ^12^, but was not a predictor of long COVID in that study. Similarly, we also observed that HAGH was not specifically a feature of long COVID, but rather a feature of SARS-CoV-2 infection as HAGH was a top protein for both COVID-19 recovered and long COVID individuals. Nevertheless, taken together this further verifies a signature of neurological alteration after SARS-CoV-2 infection. Features within this signature may prove useful in monitoring cognitive functions in affected individuals.

Long COVID has also been associated with specific neuro-inflammatory biomarkers including IL-6, TNF-α, CCL23, G-CSF ^40,41^. More recently, upregulated G-CSF and MCP-1 have been linked to brain fog ^42^. While our study corroborates prior research connecting long COVID to specific neurological and inflammatory biomarkers such as CCL23, G-CSF, and MCP-1, we found that only MCP-1 was noticeably elevated aligning with previous report ^30^. In contrast reduced levels of CCL23 and G-CSF were detected in long COVID individuals compared to healthy individuals. The role of both CCL23 and G-CSF in COVID-19 immunity remains perplexing as these proteins have been linked to both severe disease and/or COVID-19 related mortality in COVID-19 patients ^43–48^ as well as being linked to asymptomatic disease and survival ^49,50^. Similarly, unchanged or downregulated MCP-1 has also been associated with long COVID ^51,52^. Thus, the biological relevance of the association with long COVID for these proteins are likely multifaceted.

A noteworthy, and to our knowledge, unique aspect of our study was the access to paired samples that allowed analysis of inflammatory profiles after COVID-19 booster vaccination and breakthrough infections in groups of healthy, COVID-19 recovered and long COVID individuals. Using mixed-effects model, we found that before vaccination, protein levels, particularly CASP8 and HAGH, were elevated in plasma of long COVID individuals. Interestingly, the atypical protein levels of CASP8, HAGH and to some extent LAT normalised to ranges similar to those observed in the healthy and COVID-19 recovered groups after the COVID-19 booster vaccine. Whether this normalisation is a vaccine-induced effect in long COVID individuals remains to be established and could be achieved by comparing plasma protein profiles immediately before and after vaccination. Alternatively, it is possible that the underlying factors driving atypical protein expression may gradually diminish over time, as suggested by previous research ^53^. It is also possible that the observed changes in certain proteins (such as CASP-8, HAGH and LAT) correlate with the resolution of cognitive symptoms within our cohort. However, due to the limited sample size, we were unable to thoroughly investigate this potential association.

Importantly, we established that the atypical levels of HAGH and CASP8 were not exacerbated nor mirrored by re-infection. Furthermore, we showed that protein levels for SIRT2, 4E-BP1, CCL4 and STAMBP in long COVID individuals and healthy SARS-CoV-2 naïve controls were similar before vaccination. In contrast, COVID-19 recovered individuals had lower levels of these specific molecules especially for SIRT2 and 4E-BP1. The similarity in this profile between long COVID individuals and healthy individuals, and their difference from COVID-19 recovered individuals, suggests a potential lack of down regulation of these proteins in response to primary infection in long COVID individuals. This is particularly interesting to observe with 4E-BP1 and SIRT2 which both have been linked to viral replication. 4E-BP1 is associated with the PI3K/AKT/mTOR pathway and blocking this pathway can have antiviral effects. For instance, modulation of this pathway has previously demonstrated inhibition of Middle East respiratory syndrome coronavirus (MERS-CoV) replication^54^. Similarly, SIRT2 inhibition can provide broad-spectrum antiviral effects including inhibition including inhibition of coronavirus replication ^55,56^. Unaltered levels of these proteins could therefore indicate a potential dysregulation of antiviral processes typically induced to inhibit SARS-CoV-2 replication. Intriguingly the same pattern is not observed upon re-infection in long COVID individuals. Furthermore, decreased levels of both 4E-BP1 and SIRT2 in response to infection are also reflected in previously SARS-CoV-2 naïve healthy individuals. However, whether 4E-BP1 and SIRT2 detected in plasma correlates to intracellular levels and activity remains to be determined. Finally, we assessed SARS-CoV-2-specific Spike and NP IgG antibody levels in all study participants. Elevated SARS-CoV-2-specific antibody titres have previously been reported in long COVID individuals ^7,57,58^. Our group had associated these elevated titres with long COVID symptoms such as fatigue ^59^ in low transmission setting, and some studies suggest a potential role of viral persistence in driving these responses ^60,61^. Other studies have found no significant difference in antibody titres between individuals with long COVID and those who have recovered ^30,62^. It has been proposed that this may reflect long COVID individuals that follow two main patterns of antiviral immune responses. In some individuals, the response is robust, with strong cellular and humoral responses similar to those seen in people who have fully recovered and in others, there are lower antiviral responses, but still with detectable antigen-specific CD4+ T cells and/or antibodies ^63^. We observed that both Spike and NP-specific IgG antibodies were lower in the long COVID group than in the recovered group 6-9 months after primary infection. Although this observation is likely the result of multiple factors, future studies could specifically investigate antibody responses in parallel to 4E-BP1 and SIRT2 plasma concentrations in a larger cohort to further decipher the potential divergent antiviral activities in long COVID.

There are some differences and limitations in our study which may be reflected in the protein biomarkers discovered. Samples were selected based on the availability of specimens that met our long COVID criteria and had been collected at a specific timepoint after diagnosis. Additionally, for the longitudinal analysis, we were restricted by samples available for individuals that had also received a COVID-19 booster vaccination and had had a breakthrough infection while enrolled in the study. For these reasons, our sample set was also heterogeneous in terms of long COVID symptoms. Consequently, this limited the statistical power of our analyses, particularly for subgroup analyses and examining differing long COVID trajectories. This limitation means that non-significant findings may mask true associations between variables, necessitating cautious interpretation of regression estimates following booster and (re-)infection. Larger cohort sizes are needed in future studies to validate these findings. Furthermore, we lack information on the specific variant responsible for breakthrough infections. Although the timing of these breakthrough SARS-CoV-2 infections in our cohort aligns with epidemiological data indicating that most community infections at the time were due to the Omicron variant ^64,65^, definitive conclusion on breakthrough infective strains cannot be drawn. Finally, our analyses does not reveal how or whether our putative biomarker proteins are mechanistically linked to long COVID and further studies need to be conducted to explore their functional role in this context.

In conclusion, we identified several biomarkers that were substantially associated with long COVID which can be further explored to discern long COVID disease subsets. Our longitudinal analysis demonstrated that COVID-19 booster vaccination efficiently induced high levels of antigen-specific antibodies without resulting in exacerbation of atypical inflammatory and neurology-related plasma protein levels. Notably, we established that plasma profiles following SARS-CoV-2 breakthrough infection in long COVID individuals diverged from the primary infection that lacked features of previously noted dysregulation. These findings provide intriguing and valuable insight into immune response outcomes, especially to SARS-CoV-2 re-infections, in individuals with long COVID which may facilitate long COVID management strategies for this poorly understood condition.

## Data Availability

Data and code availability: To protect patient privacy, this study uses anonymised data from the COVID PROFILE study. Access to more data requires committee approval in Melbourne and will be provided by the corresponding author upon request. This paper does not report original code. This project leverages open-source R code. We have documented the specific R packages and functions used in relevant sections. Any additional information required to reanalyse the data reported in this paper is available from the lead contact upon reasonable request.

## Acknowledgements

We wish to thank Anne Hart, Maureen Ford and all members of the COVID PROFILE consortium and the study participants in the COVID PROFILE study. We also wish to thank Imadh Azeez and Dr. Lauren Smith for valuable discussion and input around data analysis. Dr. Bansal received funding from the University of Bergen, The National Graduate School in Infection Biology and Antimicrobials (or IBA) and Pasteur legatet & Thjøtta’s legat, University of Oslo, Norway [101563]. The COVID PROFILE study was supported by WHO Unity funds (2020/1085469-0), and WEHI philanthropic funds. I.M. is supported by an NHMRC Senior Research Fellowship (#1043345). This work was made possible through Victorian State Government Operational Infrastructure Support and Australian Government NHMRC IRIISS.

## Author contribution

SO and EME conceptualised the study. AB, SO, JC and EME performed the data analysis. SO, NWK and RM generated the data. AB, SO, NWK, RB and EME designed experimental work, AB, SO and EME wrote the first draft of the manuscript. RB, IM and RJC provided guidance and discussion and edited the manuscript.

**COVID PROFILE consortium members**: Emily M. Eriksson, Anne Hart, Maureen Forde, Siavash Foroughi, Nicholas Kiernan-Walker, Ramin Mazhari, Erin C. Lucas, Mai Margetts, Anthony Farchione, Dylan Sheerin, George Ashdown, Rachel Evans, Catherine Chen, Shazia Ruybal-Pesántez, Eamon Conway, Marilou H. Barrios, Jasper Cornish, Maria Edmonds, Lee M. Henneken, Lisa J. Ioannidis, Sam W. Z. Olechnowicz, Ryan B. Munnings, Joanna R. Groom, Diana S. Hansen, Rory Bowden, Anna K. Coussens, Jason A. Tye-Din, Vanessa L. Bryant, Ivo Mueller

## Declaration of interests

The authors declare no competing interests.

## Data and code availability

To protect patient privacy, this study uses anonymised data from the COVID PROFILE study. Access to more data requires committee approval in Melbourne and will be provided by the corresponding author upon request. This paper does not report original code. This project leverages open-source R code. We have documented the specific R packages and functions used in relevant sections. Any additional information required to reanalyse the data reported in this paper is available from the lead contact upon reasonable request.

## Notes

### Competing Interest Statement

The authors have declared no competing interest.

### Author Declarations

Ethics committee of the Walter and Eliza Hall Institute (#20/08) and Melbourne Health (RMH69108) gave ethical approval for this work. All subjects provided written informed consent to participate in this study, in accordance with the Declaration of Helsinki.

