## Supplementary for "Divergent inflammatory and neurology-related plasma protein profiles in individuals with long COVID following primary and breakthrough SARS-CoV-2 infections"

1. List of investigators

In addition to the authors, the following investigators contributed as part of their role in the COVID PROFILE consortium:

Siavash Foroughi^1,2^, Jason A. Tye-Din^1,2,3,4^, Anna K. Coussens^1,2,5^ and Vanessa L. Bryant^1,2,6^

Affiliations

1. Immunology Division, Walter and Eliza Hall Institute of Medical Research, Parkville, VIC 3052, Australia

2. Department of Medical Biology, The University of Melbourne, Parkville, VIC 3052, Australia

3. Department of Gastroenterology, The Royal Melbourne Hospital, Parkville, VIC 3052, Australia

4. Centre for Food & Allergy Research, Murdoch Children's Research Institute, Parkville, VIC, Australia

5. Wellcome Centre for Infectious Diseases Research in Africa, Institute of Infectious Disease and Molecular Medicine, Department of Pathology, University of Cape Town, South Africa

6. Department of Clinical Immunology and Allergy, The Royal Melbourne Hospital, Parkville, VIC 3052, Australia

7. Population Health and Immunity Division, Walter and Eliza Hall Institute of Medical Research, VIC 3052, Australia

Contributions

J.T-D and V.B contributed to cohort design; S.F designed the cohort database and managed clinical data; A.K.C assisted with study collaborative initiative and sample logistics and processing.

**Supplementary Figure 1.**

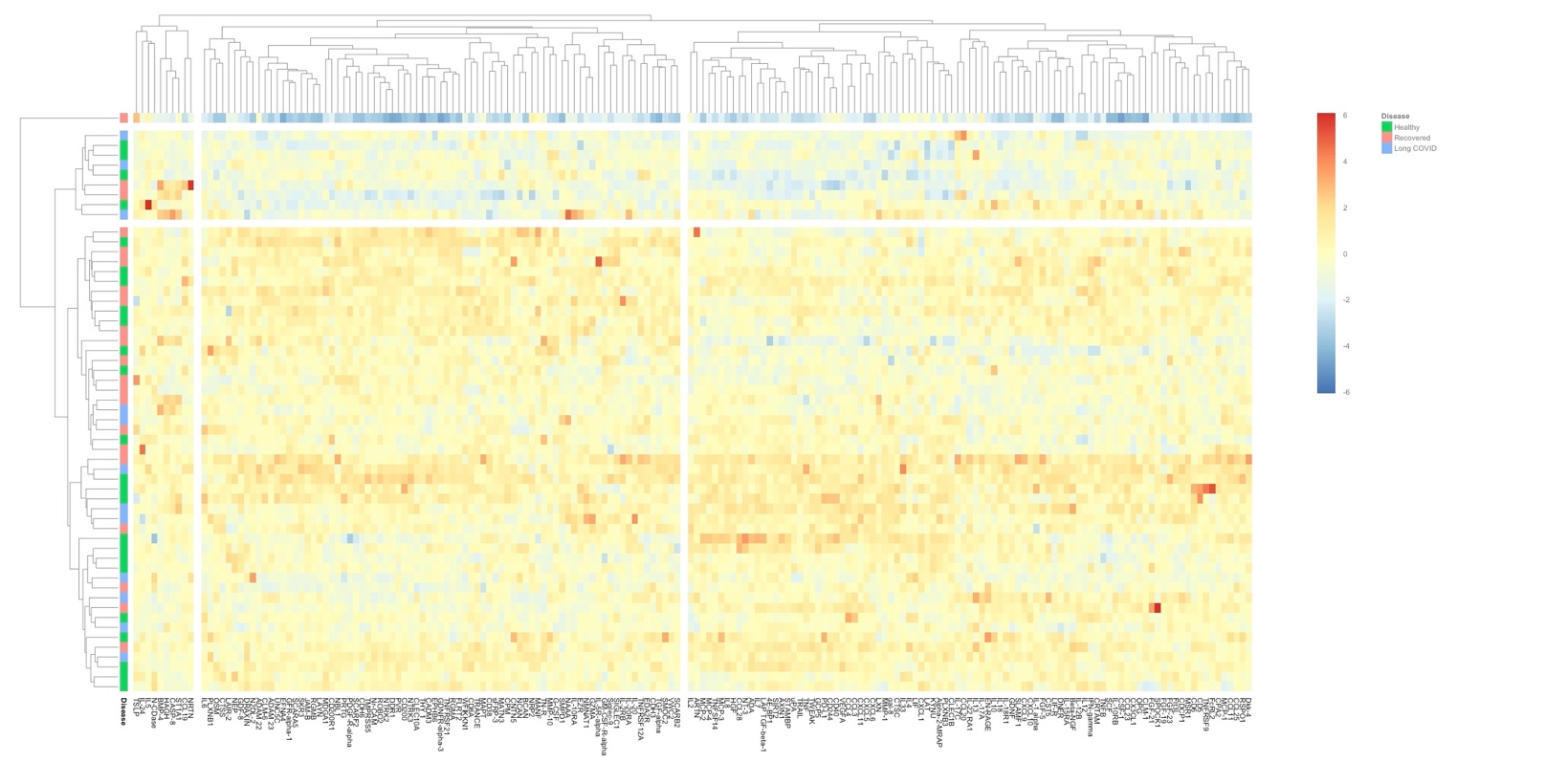

**Fig. S1**. **Olink heatmap of analyte expression for healthy, COVID-19 recovered and individuals with long COVID at 6-9 months post-infection**

This heatmap visualizes expression levels of 182 analytes in pre-vaccination samples from three groups: long COVID (blue), COVID-19 recovered (red), and healthy (green). Row-wise hierarchical clustering (cluster_rows = T) identifies groups of analytes with similar expression patterns against three groups. The dendrogram (cutree_rows = 3) displays the major clusters using olink_heatmap_plot function ^1^.

**Supplementary Figure 2.**

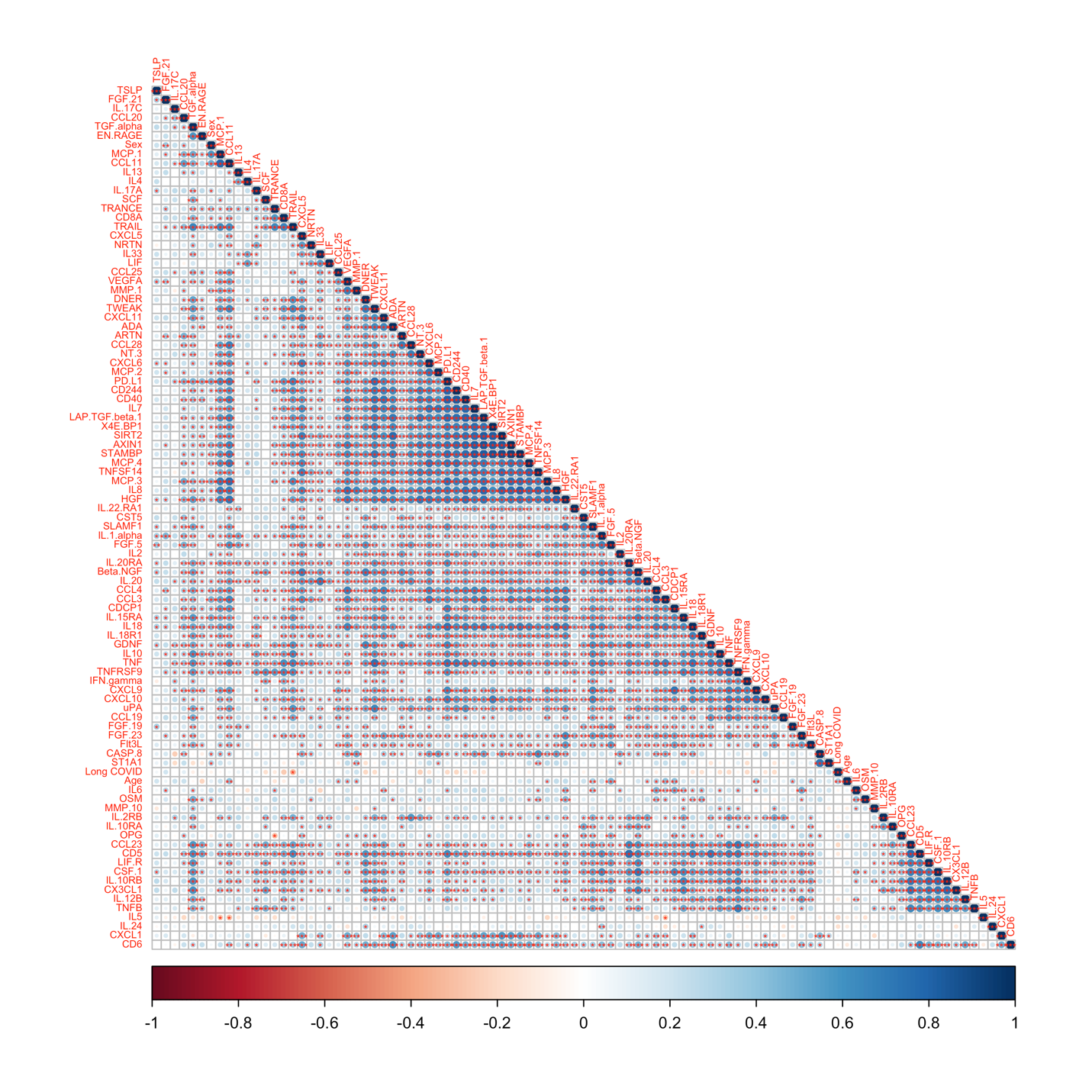

**Fig. S2 Correlation matrices: Inflammation panel analytes and long COVID.** Spearman correlation coefficients (non-parametric) between various analytes from the Olink inflammation panel. Hierarchical clustering (hclust) was used to group analytes with similar correlation patterns ^2^. Statistical significance is indicated by asterisks: *: p-value < 0.05, **: p-value < 0.01 and ***: p-value < 0.001.

**Supplementary Figure 3.**

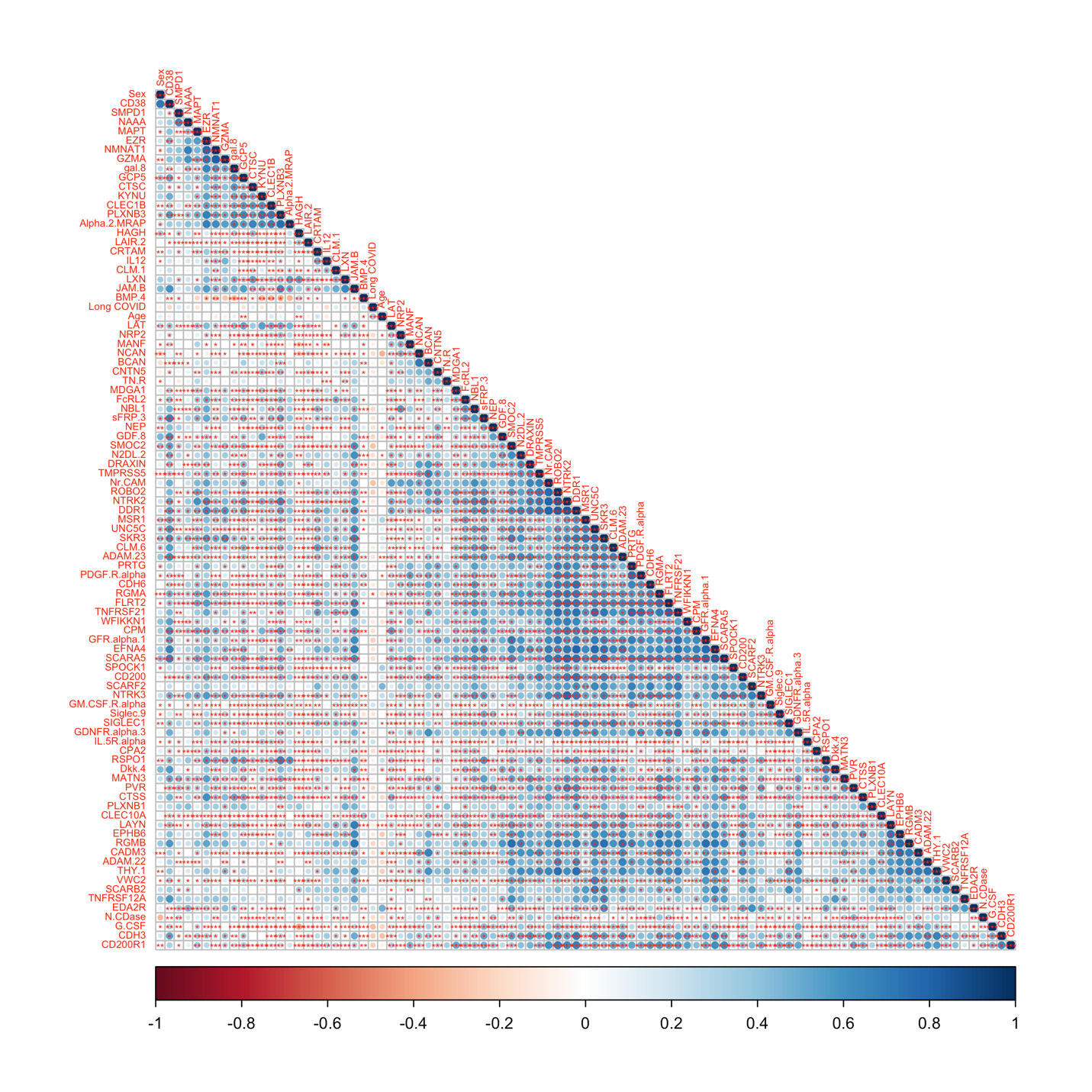

**Fig. S3 Correlation matrices: Neurology panel analytes and long COVID**. Spearman correlation coefficients (non-parametric) between various analytes from the Olink neurology panel. Hierarchical clustering (hclust) is used to group analytes with similar correlation patterns ^2^. Statistical significance is indicated by asterisks: *: p-value < 0.05, **: p-value < 0.01 and ***: p-value < 0.001.

**Supplementary Figure 4.**

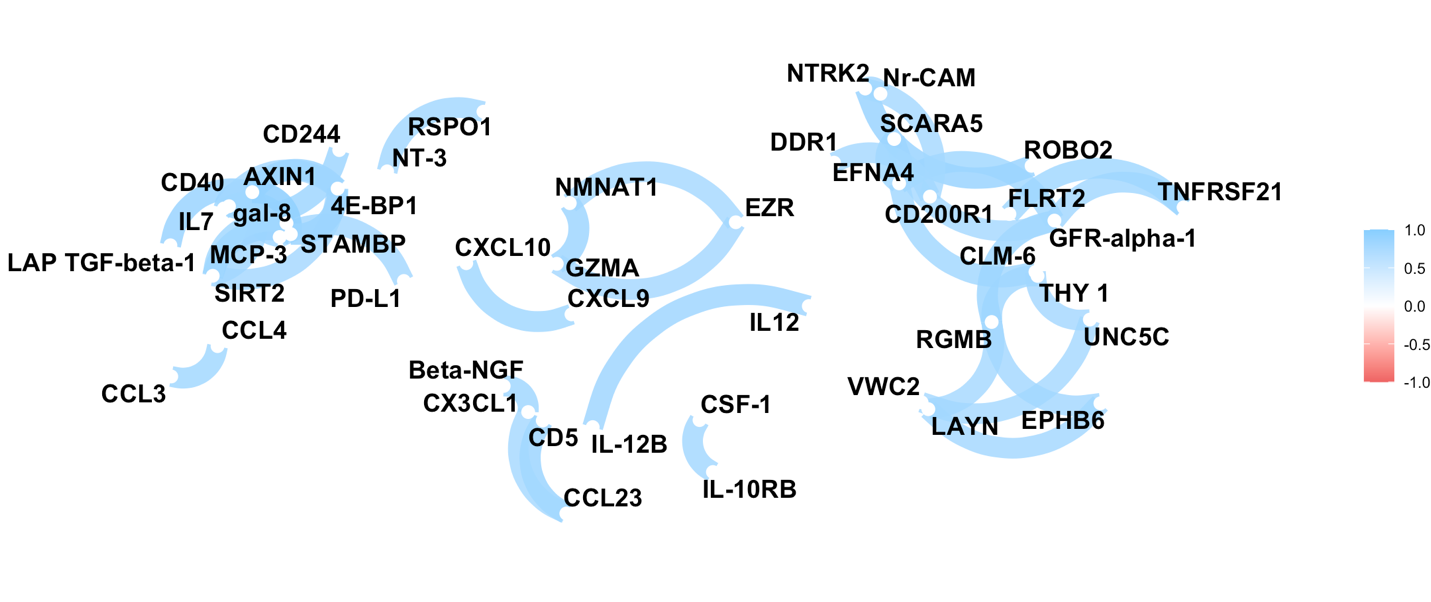

**Fig. S4. Network plot of correlated data frame with 182 analytes, COVID-19 recovered and long COVID groups.**

This figure shows a network visualization of correlations between analytes in the COVID-19 recovered and long COVID groups. The analysis used the network_plot function from the corrr package. Closer proximity: Indicates stronger correlations between assays with blue for positive correlation and only correlations with an absolute value greater than 0.8 (stronger correlations) are shown (using min_cor = .8). The positioning of assays is determined by multidimensional clustering, grouping assays with similar correlation patterns {Kuhn, 2022 #72

**Supplementary Figure 5.**

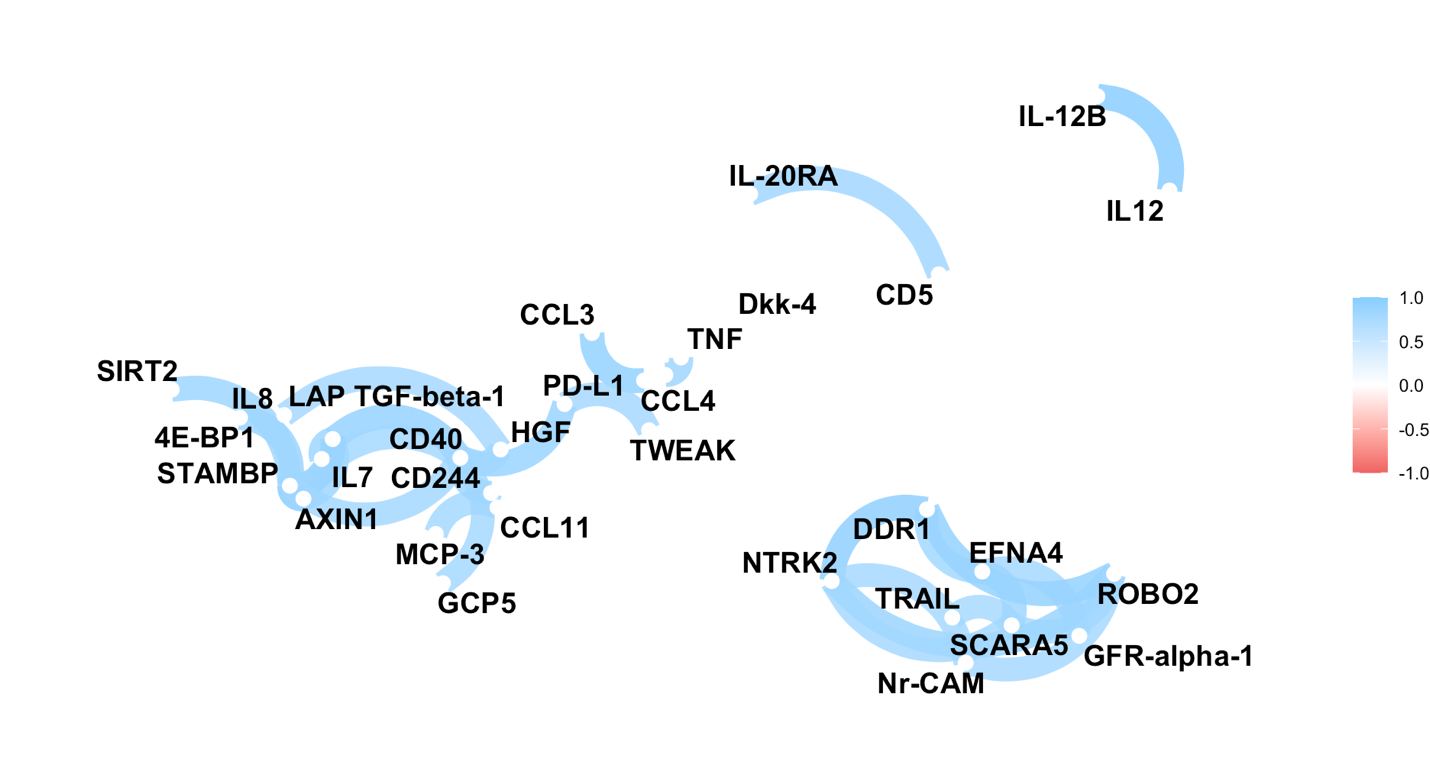

**Fig. S5 Network plot of correlated data frame with 182 analytes, healthy and long COVID group**

This figure shows a network visualization of correlations between analytes in the healthy and long COVID groups. The analysis used the network_plot function from the corrr package. Closer proximity: Indicates stronger correlations between assays with blue for positive correlation and only correlations with an absolute value greater than 0.8 (stronger correlations) are shown (using min_cor = .8). The positioning of assays is determined by multidimensional clustering, grouping assays with similar correlation patterns {Kuhn, 2022 #72}.

**Supplementary Figure 6.**

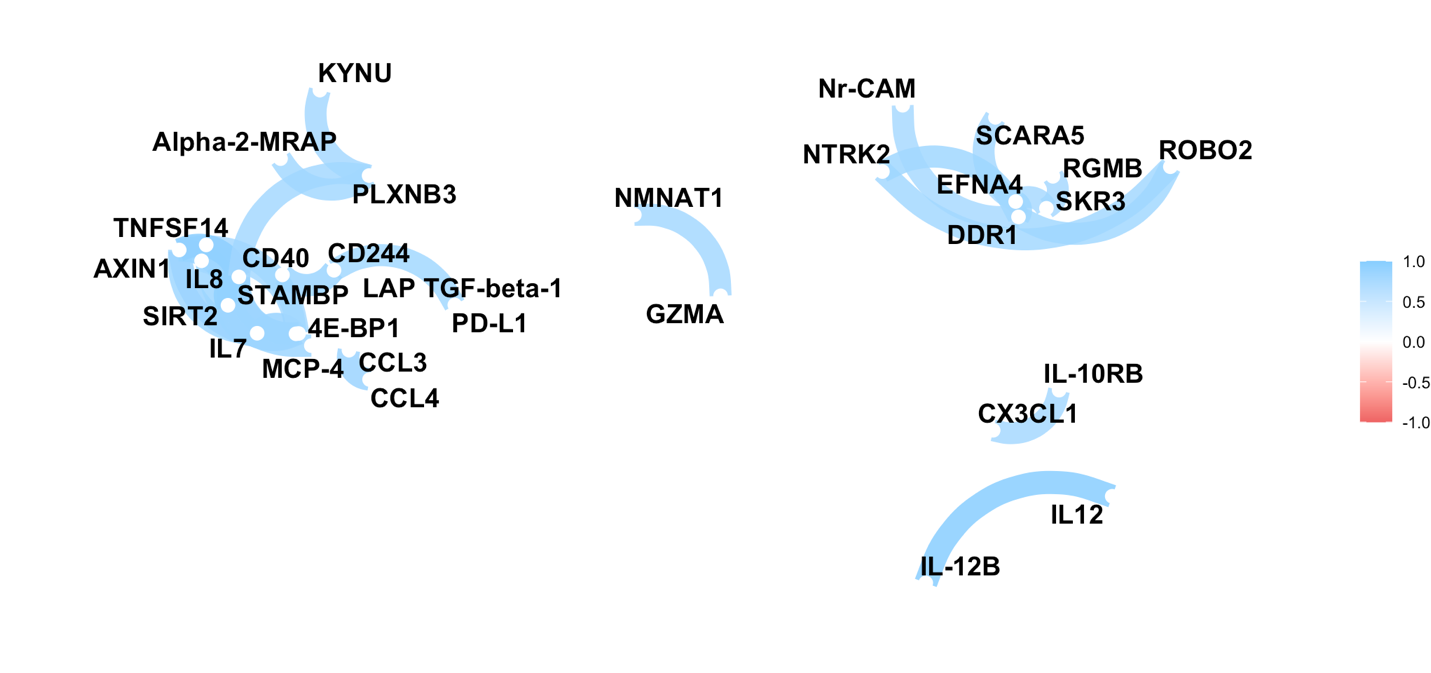

**Fig. S6 Network plot of correlated data frame with 182 analytes, healthy and COVID-19 recovered groups.**

This figure shows a network visualization of correlations between analytes in the recovered and healthy groups. The analysis used the network_plot function from the corrr package. Closer proximity: Indicates stronger correlations between assays with blue for positive correlation and only correlations with an absolute value greater than 0.8 (stronger correlations) are shown (using min_cor = .8). The positioning of assays is determined by multidimensional clustering, grouping assays with similar correlation patterns ^3^.

**Supplementary Figure 7.**

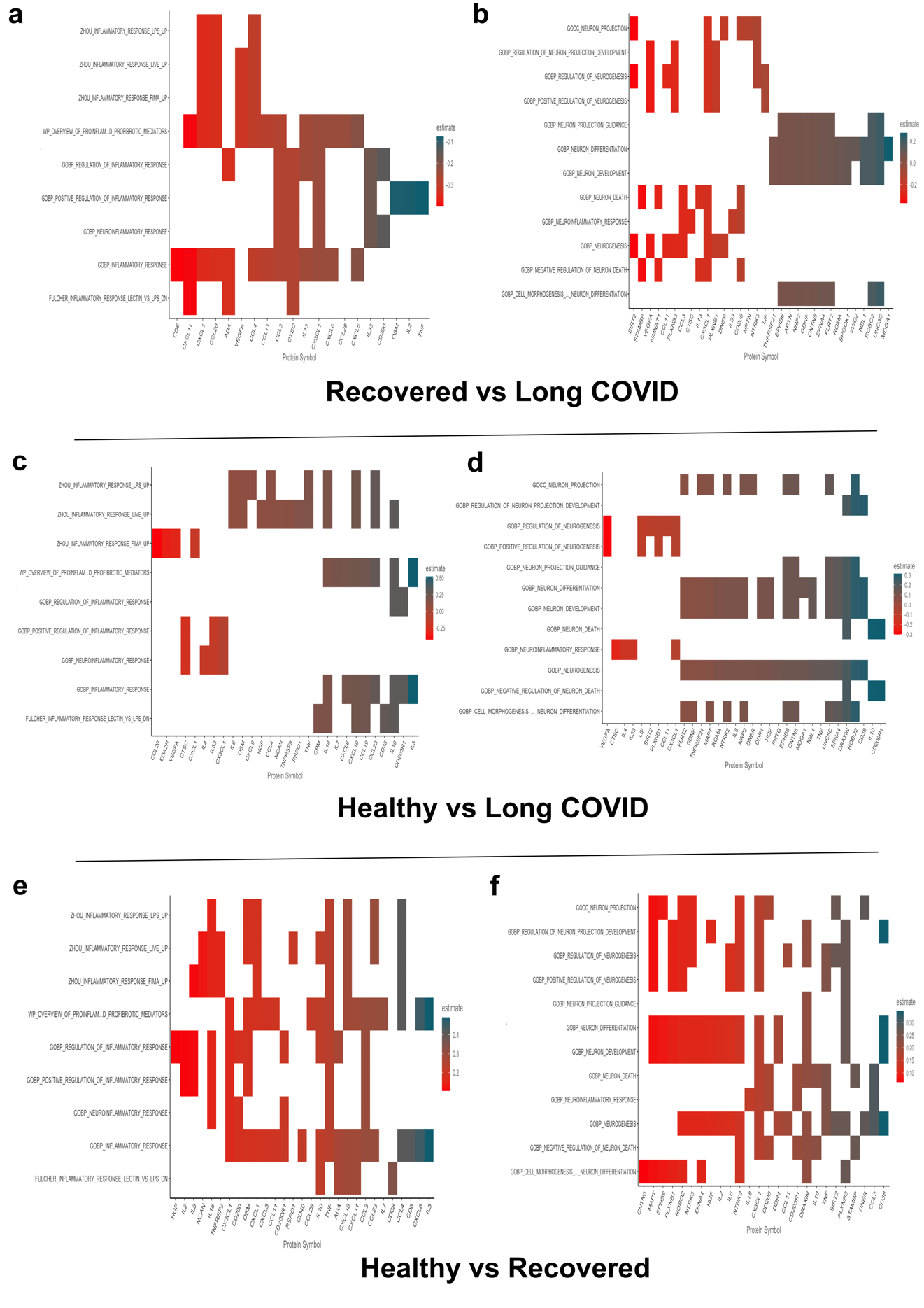

**Fig. S7 Olink gene set enrichment analysis (GSEA) for inflammation and neurology pathways at 6-9 months post-infection.**

The enrichment of inflammation (a,c, e) and neurology (b,d,f) pathways was explored in three groups (COVID-19 recovered vs. long COVID; top panel, healthy controls vs. long COVID; middle panel, and healthy controls vs. COVID-19 recovered; bottom panel). The analysis was performed in R using the following functions: a) olink_ttest: This function performs t-tests to compare the expression levels of analytes between the groups. b) olink_pathway_enrichment: This function utilizes Gene Set Enrichment Analysis (GSEA) to identify pathways (groups of proteins with related functions) that are significantly enriched in one group compared to another. Only inflammation and neurology pathways are shown due to the panel selection. Pathway analysis reflects our focus and may miss pathways outside our focused specific biological areas ^1,4^.

1	Nevola K *et al.* _OlinkAnalyze: Facilitate Analysis of Proteomic Data from Olink_. R package version 3.6.2,. (2024).

2	Taiyun, W. & Viliam, S. R package 'corrplot': Visualization of a Correlation Matrix (Version 0.92). (2021).

3	Kuhn, M., Jackson, S. & Cimentada, J. Correlations in R_. R package version 0.4.4,. (2022).

4	Nevola K, S. M., Guess J, Forsberg S, Cambronero C, Pucholt P, Zhang B, Sheikhi M, Diamanti K, Kar A, Conze L OlinkAnalyze: Facilitate Analysis of Proteomic Data from Olink_. R package version 3.6.2. (2024).
